# Integrative multi-ethnic Mendelian randomisation identifies tissue-specific causal genes for Coronary Artery Disease and interactions with post-acute Covid

**DOI:** 10.1101/2024.10.11.24315315

**Authors:** Rachel Jaros, Justin M. O’Sullivan

**Affiliations:** The Liggins Institute, The University of Auckland, Auckland, Private Bag 92019, New Zealand; The Maurice Wilkins Centre, The University of Auckland, Auckland, Private Bag 92019, New Zealand; MRC Lifecourse Epidemiology Unit, University of Southampton, Southampton, SO16 6YD, United Kingdom; Singapore Institute for Clinical Sciences, Agency for Science Technology and Research, Singapore

## Abstract

Coronary artery disease (CAD) is highly heritable and remains the leading cause of mortality worldwide. Understanding the genetic and mechanistic underpinnings of CAD is crucial for early risk assessment and intervention. We conducted a transcriptome-wide Mendelian randomisation (MR) study, utilising unbiased tissue-specific gene regulatory networks, to identify genes causally associated with CAD in European and East Asian populations. We identified 291 tissue and ancestry-specific genes implicated in CAD, including 98 novel protein-coding genes across coronary artery, whole blood, and lung tissues. Genes involved in epigenetic processes (eg *PAXBP* and *KIAA0232*) causally associated with CAD. Moreover, we identified genes related to the ubiquitin-proteasome system in the coronary artery and kinase signalling in the lung, as being causally related to CAD. The integration of protein interaction networks identified causal connections between CAD and HDL cholesterol levels, providing novel insights into CAD mechanisms, and potential actionable targets for people with this risk profile. The results also provide intriguing insights into the link between SARS-CoV-2 and CAD, unveiling mechanisms that may underlie the increased risk of cardiovascular disease following SARS-CoV-2 infection. The causal mechanisms we identified emphasise the tissue-agnostic and ancestrally unique pathways that underscore the complex interplay between CAD development, metabolic disturbances, and the immune system. Collectively, our results provide valuable insights into CAD pathogenesis and potential therapeutic targets.

## INTRODUCTION

The prevention and treatment of coronary artery disease (CAD) has progressed significantly over the course of the last 50 years. However, CAD remains a leading cause of death worldwide^1^. Genetics are estimated to account for 40% to 60% of the risk of developing CAD. To date, GWAS have identified 150 genetic risk loci as being associated with CAD^2–6^. These loci are predominantly autosomal and have been linked to well-known disease risk factors (eg lipids and blood pressure)^7,8^. Heritability is more pronounced in younger individuals and those with a family history of premature CAD^9^. Notably, distinct CAD genetic risk factors have been identified in East Asian populations^5,6^. However, some European susceptibility loci are also significant for East Asian populations^2–4,10,11^. The existence of unique and shared loci between ancestral groups indicates the potential for a core CAD causal network and additional networks of causal genes that interact with environment specific factors. As such, it is necessary to characterise the between- and within-ancestral group genetic causes of CAD to improve risk prediction and the development of novel therapies.

CAD is a multi-tissue disease whose development begins in adolescence. However, little is known about the mechanisms through which these susceptibility loci impact CAD, their interactions in different tissues and in different ancestral populations. The strong correlation between the odds ratios (ORs; risk) for SNPs associated with CAD and 27 other diseases across European and East Asian samples^3^ indicates the potential for comorbidities to be genetically linked to CAD. Improving our understanding of comorbidities is essential as the pace of disease development is intricately associated with tissues involved in metabolic disturbances^12^, the immune system^13^ and gene x environment interactions.

Systems genetic approaches use networks of gene regulation and protein-interactions to predict how genetic variants impact disease risk^14–20^. Our approach integrates GWAS with functional genomic datasets (ie genome structure, expression quantitative expression trait loci (eQTL), and protein interaction networks^21–23^ through a de novo, 2-sample Mendelian randomisation. This enables identification of putatively causal CAD genes for use as seeds in a subsequent protein interaction network analyses. In so doing, we can identify not only the causal genes but biological pathways that connect CAD to comorbidities. Extending this ‘causal’ centred network to different ancestral populations will further enhance our ability to predict individual CAD risk and identify effective therapeutic strategies^20^.

In this study, we used gene regulatory networks (GRNs) from the coronary artery, lung and whole blood to identify tissue-specific causal genes for CAD across European and East Asian populations. Subsequent network analyses^24–27^ identified how the 98 putative causal CAD genes interact with CAD comorbidities and associated traits (eg response to SARS-CoV-2 infection). Our results link CAD causal genes with HDL cholesterol metabolism. Finally, we propose a potential mechanism that explains how sustained latent asymptomatic CAD conditions arise following SARS-CoV-2 infection.

## RESULTS

### Mendelian randomisation identifies tissue-specific and ancestrally divergent causal genes for coronary artery disease

Similarity in CAD heritability across ancestral populations (i.e. East Asian, European, African, and Indigenous American) indicates the existence of a consistent genetic influence on disease risk relative to non-genetic factors across these populations^6^. Consistent with this, tissue-specific gene regulatory modules associated with CAD have been identified^28–30^. Here, we combined a system genetics approach^16,17^, with Mendelian randomisation (MR)^31,32^ to identify CAD causal genes within biological networks. We applied two-sample MR^31,32^ using genes (exposures) from lung^21^, blood^33,34^, and coronary artery gene regulatory networks (GRNs; see methods) without *a priori* assumptions.

The GRN networks have been previously described^24–27^. Briefly, the GRNs were generated by assessing all known human SNPs for spatial interactions with genes from tissue/cell-specific chromatin conformation (ie Hi-C) data, then parsing these SNP-gene pairs through GTEx for eQTL activity within the same tissues. In order to be included in the MR, an eQTL variant must (i) exhibit strong associations with the exposure, (ii) be associated with the outcome through the tested exposure only, and (iii) have no associations with any potential confounders that affect the outcome. Genetic variants meeting those assumptions are termed instrumental variables (IVs). Therefore, we filtered each GRN for eQTLs with a *p* < 1 x 10^-5^ and performed LD clumping (r^2^ < 0.001, 10Mb away with a European population reference). The resulting spatial-eQTLs (sceQTLs) were used as IVs for MR. The lung GRN consisted of 15,532 genes regulated by 732,000 sceQTLs, the whole blood GRN consisted of 14,556 genes regulated by 720,675 sceQTLs and the coronary artery GRN consisted of 6,542 genes regulated by 144,399 sceQTLs (Figure 1; *p* < 1 x 10^-5^ and FDR < 0.05; see methods). From the GRNs, 53.6% (n = 8,336) lung genes, 85.1% (n = 5,571) coronary artery genes, and 52.9% (n = 7,708) blood genes could only be instrumented using a single sceQTL, and all MR estimates derived from these genes are based on the Wald ratio method^35^. MR estimates derived on the genes with multiple sceQTLs are based on the fixed-effects, inverse-variance-weighted (IVW-fe) method^35^. No causal genes were identified using the IVW-fe method.

**Figure 1.**
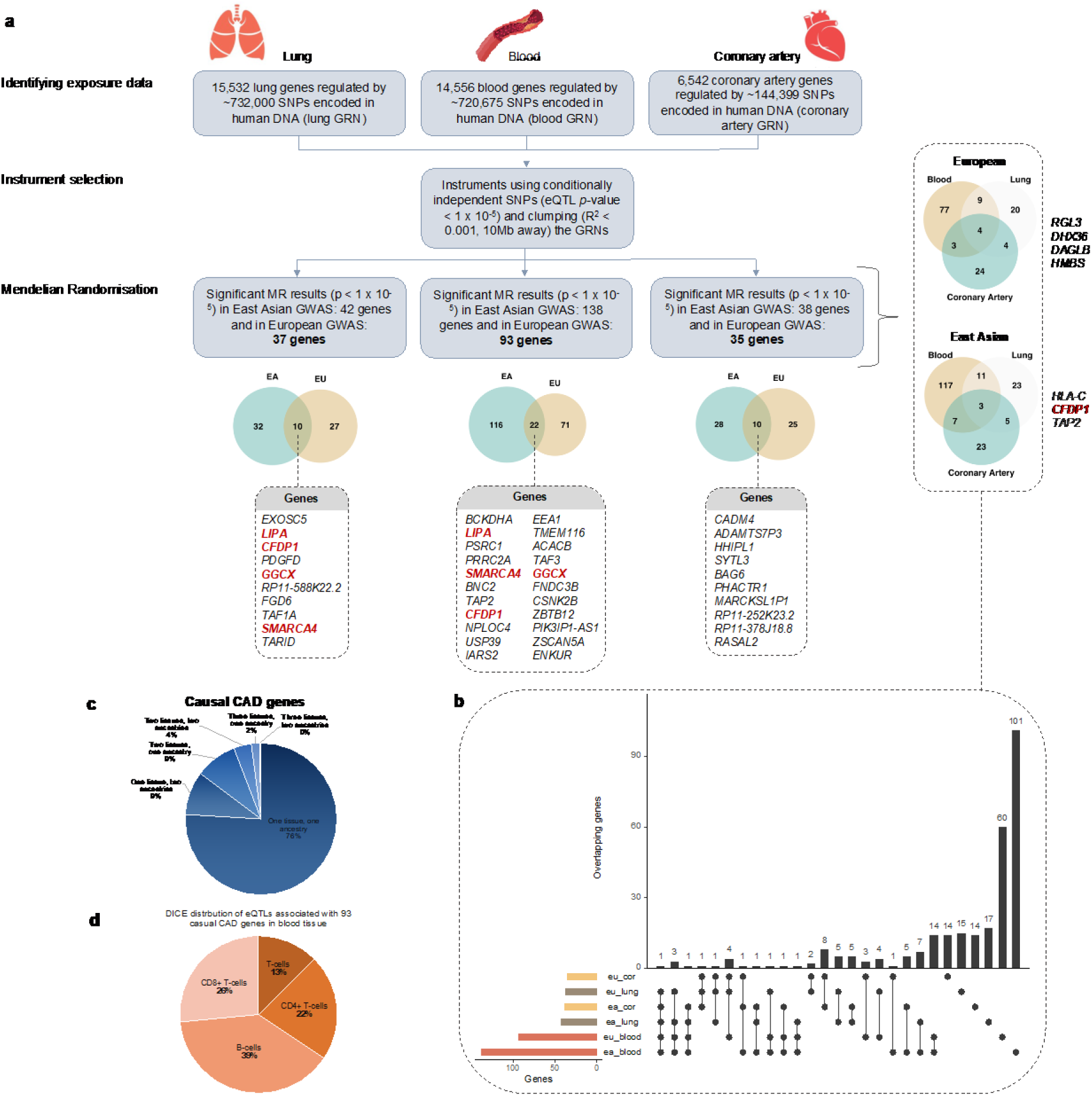
Multi-tissue causal analysis and gene regulatory networks identify ancestry-specific genes that are causal for coronary artery disease. (a) 2-sample MR identified tissue-specific CAD causal genes that were shared, or unique between the European and East Asian ancestral populations. 2-sample MR was conducted using tissue-specific spatially-constrained eQTLs (sceQTLs) as the instrumental variables (IVs) and CAD GWAS SNPs fromEuropean^6^ and East Asian^5^ populations as the outcomes (see methods), without *a priori* assumptions. The genes in red are shared. b) CAD causal genes are typically tissue-specific. eu_cor, European coronary; eu_lung, European lung; ea_cor, East Asian coronary; ea_lung, East Asian lung; eu_blood, European Blood; ea_blood, East Asian blood. c) CAD causal genes are typically Ancestry-specific. d) sceQTLs for 93 causal genes are present within T and B cells in the Database for Immune cell eQTls.

To identify ancestry- and tissue-specific *causal* genes associated with CAD we incorporated CAD GWAS data (outcome) from two distinct ancestral groups, East Asian (9,413:203040 case:control; Japanese individuals)^5^ and Europeans (190,493:582,775 case:control)^6^ from the IEU open GWAS project^36^ catalogue with the GRNs.

There is no overalp between the individuals the GWAS^5,6,36^ or GTEx study (GTEx v 8.0)^37^, as they were independent, and have not been merged. MR identified 42 and 37 genes, 138 and 93 genes, 38 and 35 genes from lung, blood and coronary artery- GRNs (East Asian and European), respectively (*p*-value < 1 x 10^-5^; Figure 1; Supplementary figure 1-3; Supplementary table 1). 152 of the 291 genes had been previously significantly associated with CAD in the Cardiovascular Disease Knowledge Portal^38^ (Supplementary table 2).

Among the novel causal genes we identified (n = 139), 98 were protein-coding, and the remaining 41 were either long non-coding RNAs, antisense RNAs, pseudogenes, or micro-RNAs. In Europeans, proportionally (1.2%) more of the causal gene set from the blood-GRN (n = 93) is associated with CAD than observed in the lung (n = 37, 0.44%), or coronary artery (n = 35, 0.62%) tissue GRNs, suggestive of a greater role for the immune system in CAD (Figure 1a) since the the cellular composition of the blood-GRN is predominantly comprised of B- and T-cell populations^39^ (Figure 1d).

To determine whether the causal blood get set was enriched in clinical tissues associated with CAD, gene set enrichment analysis^40,41^ using RNA-seq data from coronary artery plaque samples of patients with stable and advanced coronary artery disease^42^ as performed. This showed an upregulation of the 93 European causal blood genes in advanced disease, and not in stable disease. However, there was no significant enrichment (*p* = 0.164). This is likely due to the cellular composition of the plaque samples, which predominantly contain fibromyocytes, macrophages and endothelial cells^42^.

Ancestry-specific causal genes were identified for East Asian and European populations (Figure 1). 221 causal genes representing 76% of the causal gene set (Figure 1b and c; Supplementary table 1) were tissue and ancestry-specific. Four genes (*RGL3*, *DHX36*, *DAGLB*, and *HMBS*) within the European ancestry and three within East Asians (*HLA-C*, *CFDP1* and *TAP2*), 2% of the causal genes (Figure 1c), were shared across all three tested tissues (Figure 1a; Supplementary table 1). No genes overlapped across all tissues and both ancestries (Figure 1b). Expression of each of the 7 genes, that were shared across the three tissues, was associated with a unique sceQTL (Figure 2a). Of these 7 genes, four decrease and three increase the odds ratio (OR) for CAD. For example, decreased expression of *HLA-C* (East Asian) was associated with an increased CAD OR. By contrast, increased expression of *DAGLB* (European) and *CFDP1* (East Asian) was associated with a decreased CAD OR.

**Figure 2.**
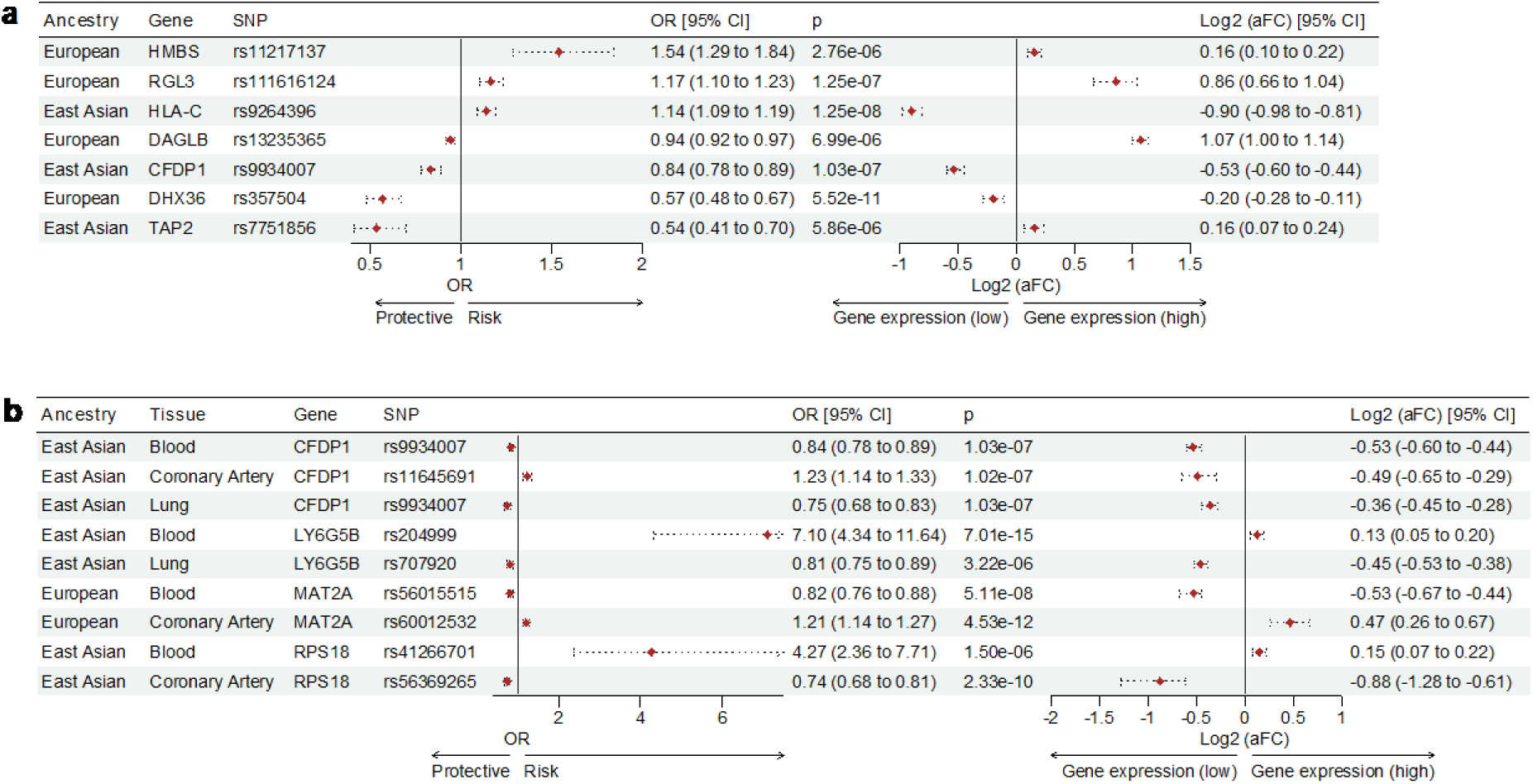
MR identified seven CAD causal genes that were shared across all three tested tissues and had distinct expression changes. Forest plots show genes identified as having significant (*p* < 1 x 10^-5^) causal effects on CAD that are ancestrally divergent but shared across the three tested tissues (see Figure 1a). The OR of causal CAD genes per 1-SD change in gene expression in the three tested tissues. The allele-specific gene expression changes (aFC) associated with the eQTLs (SNP) in the three tested tissues. 95% confidence intervals are represented by the horizontal dotted lines. (a) Seven genes were identified as being significant for CAD in all three tissues tested (b) CAD causal genes that had tissue-specific ORs consistent with both risk and protection. The forest plot shows the switch in OR between blood, coronary artery and lung (e.g. *LY6G5B* is a risk gene for CAD in blood and protective for CAD in the lung). OR, Odds ratio; aFC, allelic fold change.

We identified four causal CAD-genes (3 East Asian; 1 European), instrumented by a single IV, that have tissue-specific ORs consistent with both risk and protection (Figure 2b). In East Asians, *LY6G5B* transcript levels in whole blood were associated with rs204999 and a high risk for CAD (OR = 7.10 [4.34 – 11.64, 95% CI]). By contrast, *LY6G5B* transcript levels associated with rs707920 in lung where the OR was consistent with a small but protective effect against CAD (OR = 0.81 [0.75 – 0.89, 95% CI]). Similarly, *RPS18* transcript levels associated with rs41266701 in whole blood where the OR was consistent with high risk (OR = 4.27 [2.36 – 7.71, 95% CI]) for CAD. Notably, *RPS18* transcript levels associated with rs56269265 in the coronary artery and were protective against CAD (OR = 0.74 [0.68 – 0.81, 95% CI]). Similarly, transcript levels for *CFDP1* associated with rs11645691 in the coronary artery where it was high risk (OR = 1.23 [1.14 – 1.33, 95% CI]). By contrast, transcript levels for the *CFDP1* associated with rs9934007 in the lung and rs9934007 in whole blood where the gene was associated with a lower risk of developing CAD (Figure 2b). Notably, *CFPD1* associated with rs9934007 was also causally associated in whole blood and lung in Europeans (Supplementary table 1). Only *MAT2A* exhibited eQTL-specific switching from CAD risk to protection (coronary artery and whole blood, respectively) in Europeans (Figure 2b).

We hypothesised that alterations in relative isoform levels were associated with the eQTL-gene transcription changes, in the tissues in which they switched from being CAD risk to protection (Figure 2). Gene isoform relative ratios changed between tissues for the four causal genes that switched from risk to protection (Figure 2b). For example, *RPS18* isoform expression in whole blood was 0.184 ± 0.06 (average ± SD) of the expression observed in coronary artery, in samples from GTEx^37^. Similarly, the expression of the *MAT2A* isoforms in coronary artery was 47.42 ± 18 (average ± SD) that observed in whole blood samples from GTEx^37^. The variation in *MAT2A* isoform expression was driven by ENST00000306434.7 whose coronary:blood ratio was only 15.8. *CFDP1* isoform expression ratio changed on average by 0.815 ± 0.33 coronary artery:lung and 8.35 ± 5.8 coronary artery:whole blood. Expression of three *CFDP1* isoforms (ENST00000570103.5, ENST00000564286.1, and ENST00000570279.1) was not detected in whole blood. Finally, *LY6G5B* isoform expression whole blood:lung ratios averaged 0.721 ± 1.018368042, with the variation being due to an increase in ENST00000471088.1 in while blood (Supplementary table 3).

We next assessed the levels of individual transcripts for *RPS18*, *MAT2A*, *CFPD1* and *LY6G5B*. *LY6G5B* transcript ENST00000471088.1 levels increase 1.8 times when comparing isoform expression patterns in whole blood and lung. By contrast, the relative ratio of ENST00000471088.1 (ratio = 0.48, *p* = 0.33; Supplementary table 3) when compared to the main *LY6G5B* transcript (ENST00000409525.1) in whole blood is approximately ten fold that observed when comparing expression levels of these isoforms in lung (ratio=0.039). Notably, ENST00000409525.1 does not contain an open reading frame (ORF) and is not predicted to produce a protein^43^.

*CFDP1* is essential for cardiac development in zebrafish^44^. *CFDP1* was identified as being causal in all three tested tissues, but with tissue-specific ORs indicating risk (coronary) and protection (blood and lung; Figure 2b). *CFDP1* is expressed as eight isoforms in the coronary artery (Supplementary table 3). Notably, the ENST00000564286.1 isoform transcript level is 1.2 times greater than observed in the lung (*p* = 0.14) and whole blood (*p* = 0.02) and exhibits a increase in expression overall (ΔTPM= 0.085, Supplementary table 3). No protein-coding sequence (CDS) has been defined for isoform ENST00000564286.1^44^. However, the main protein coding transcript (ENST00000283882.3) is 8.4 times more highly expressed in the coronary artery, and is associated with increased risk when compared to whole blood. This increase in expression may explain the tissue-specific effect of *CFDP1* being risk in the coronary artery and protective in whole blood and lung.

*MAT2A* is associated with risk in the coronary artery and protection in whole blood (Figure 2b). Expression of *MAT2A* isoforms ENST00000409017.1 and ENST00000481412.5 increased relative to the main isoform (ENST00000306434.7) in whole blood and the coronary artery. The transcript expression change for the isoforms expressed in the coronary artery, when compared to whole blood, ranged from 15.85 to 68.29 (*p* = 1.05 x 10^-5^). For example, transcript ENST00000409017.1 which is protein coding for MAT2A increases by 40.98^43^ (Supplementary table 3). Therefore, it is possible that increases in the expression of this isoform, or changes relative to the other isoforms, may be directly linked to increased risk of CAD.

Two isoforms of *RPS18* (ie. ENST00000474626.5 and ENST00000496813.5) increase their levels relative to ENST00000439602.6 in whole blood when compared to the coronary artery (Figure 2b; Supplementary table 3). Both the ENST00000474626.5 and ENST00000496813.5 isoforms are biotyped to have a retained intron relative to other coding, transcripts of *RPS18*^43^.

We hypothesised that the causal genes within the lung, whole blood and coronary artery tissues contribute to the total genetic burden by forming interactions and entire functional subnetworks with other genes that are expressed in a tissue-specific manner. Pathway analyses were performed on the complete set of causal genes that was identified within each tissue/ancestral group (Supplementary table 4). g:Profiler^45^ identified significant (FDR *p* < 0.05) enrichment in 32, 56, 13 pathways within the East Asian blood, coronary artery and lung, respectively. Similarly, 26, 1 and 29 pathways were enriched within the European blood, coronary artery and lung causal gene sets, respectively. The enriched pathways included triglyceride lipase activity (*p* = 2.03 x 10^-3^) and peptide antigen binding (*p* = 9.17 x 10^-4^). The European lung causal gene set uniquely featured pathways known to be involved in CAD including phospholipid homeostasis (*p* = 2.63 x 10^-3^), cholesterol metabolism (p = 3.00 x 10^-4^), metabolic pathway of LDL, HDL and TG, including diseases (*p* = 5.61 x 10^-4^) and atherosclerosis (p = 0.024) (Supplementary table 4).

### Traits that co-occur with CAD are pleiotropic and involved in metabolic disturbances

Metabolic disturbances (dyslipidaemias, diabetes, overweight) are of particular importance to the development of CAD^12^. Notably, immune responses participate in every phase of atherosclerosis^13^. Since we identified a higher proportion of genes within whole blood (Figure 1b and c) as being causal for CAD, we used the whole blood GRN^33,34^ and experimentally validated protein interactions^46^ to assess the co- occurrence of traits with CAD. The chromatin interaction data, used to assess the whole blood eQTLs for the 93 causal European ancestry genes, was comprised of eQTLs from T-cells (61%) and B-cells (39%; Figure 1d).

We created a ‘‘CAD-associated network’ (Figure 3a and b) and CAD-causal network’ (Figure 3a and c) by seeding protein interaction networks with genes (protein_0_) whose transcript levels are correlated with CAD-associated GWAS variants or with causals genes from the European population^6^ within the whole-blood GRN^33,34^. Bootstrapping (n = 1,000; p ≤ 0.05) identified metabolic disturbances as being pleiotropic with the genes/proteins at level 0 (Figure 3) in both the CAD-causal and CAD-associated networks.

**Figure 3.**
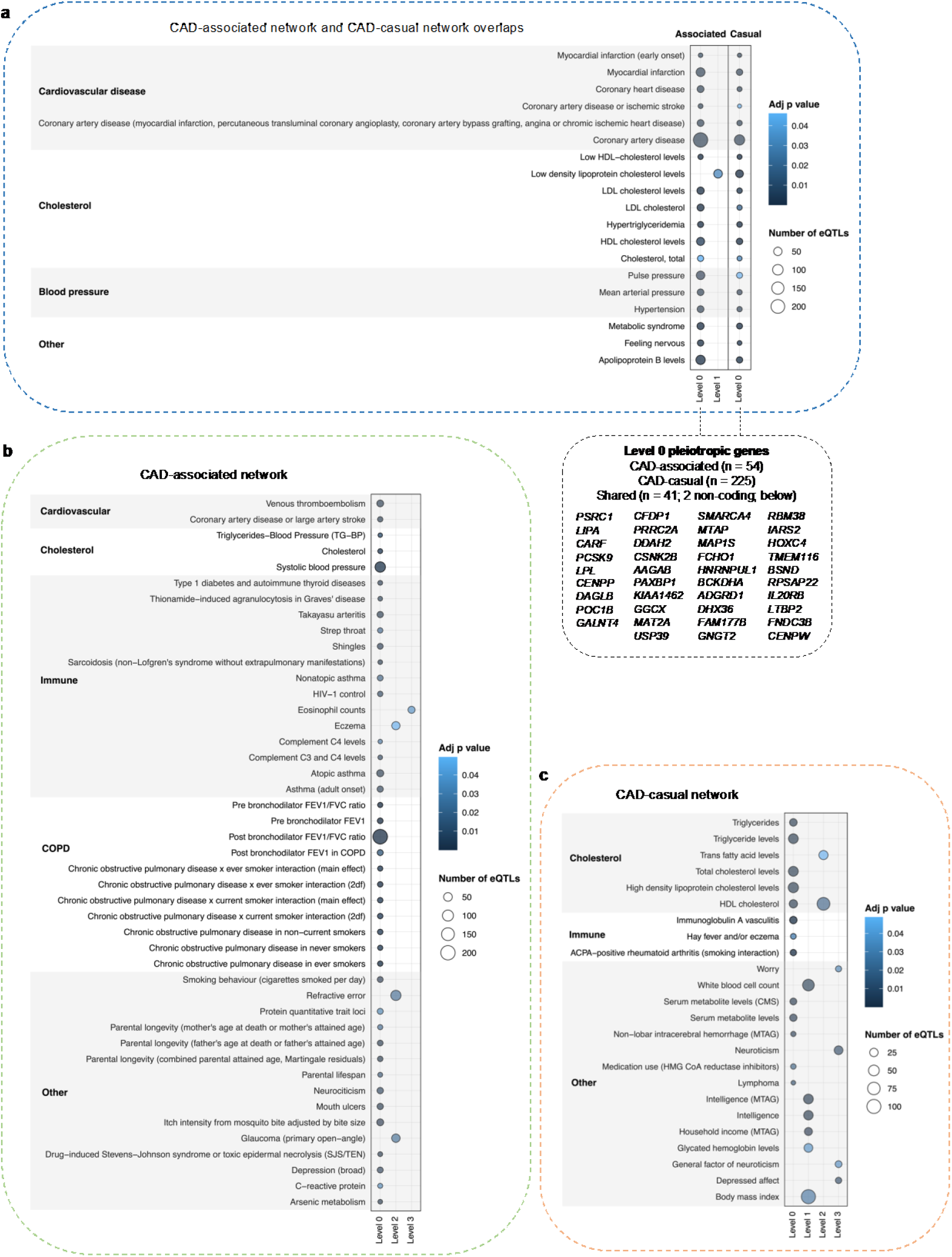
Causal and associated networks converge on the same comorbidities that co-occur with CAD. Traits that co-occur with CAD were identified using protein interaction networks generated from CAD causal genes and associated GWAS SNPs. Trait-associated loci enriched within (a) Shared CAD-causal and associated networks, (b) CAD-associated only network and (c) CAD-causal only network. Protein-protein interaction network analyses were seeded (level 0) using either: 1) CAD causal genes identified by MR in whole blood in Europeans (Figure 1b; n = 93); or 2) genes whose transcript levels correlate with CAD-associated GWAS variants (Supplementary table 5). Networks were expanded to level 4 by querying the PROPER-Seq^46^ database for interacting proteins (methods). The blood GRN was queried to obtain all known sceQTLs for each protein within the expanded networks. eQTLs from each level were then tested for enrichment (hypergeometric test) within the GWAS Catalog ^47^ to identify associated phenotypes. Traits identified at L0 of the CAD-causal network are inferred to be comorbid traits of CAD, whereas those identified at L0 of the CAD-associated network are CAD-associated traits. Target genes of the regulatory loci associated with L0 traits within the CAD-causal and associated networks are highlighted in the green box. Genes listed are shared between the two networks.

44 traits were significantly enriched (adj *p* < 0.05) within the CAD-causal network (Figure 3a and c; Supplementary table 6a). Six of the traits were CAD or proxy-CAD (first degree relative, e.g. coronary heart disease and myocardial infarction). This is consistent with the identification of the CAD causal genes (n = 93) as regulatory sceQTLs for these genes have previously been associated with CAD by GWAS. Notably, 13 of the co-occurring traits within the causal network are associated with cholesterol, consistent with well-established epidemiological and genetic observations that cholesterol traits are causal for CAD when tested using MR^48^. The cholesterol traits we identified at level 2 (i.e. trans fatty acid levels and HDL cholesterol levels; Figure 3c) support our hypothesis that modifiers of causal genes exist. For example, the HDL cholesterol trait at level 2 was associated with a set of 80 SNPs regulating 60 genes (Supplementary table 7c-d), five of which (*ZNF664, MAP3K11, UBASH3B, MARCH2, R3HDM2*) were on level 0 and causal for CAD using two-sample MR (Wald ratio; *p* < 0.001; Supplementary figure 4; Supplementary table 7a). *ZNF664, UBASH3B, MARCH2, R3HDM2* have not been linked to CAD previously.

Regulatory loci associated with level 0 traits in the causal network directly modulate the CAD causal genes. Therefore, it can be inferred that these traits are ‘comorbid’ with CAD and they may contribute to worsen the disease trajectory (Figure 3). By contrast, the regulatory loci associated with traits spanning levels 1 - 3 within the CAD- causal network influence the causal genes by acting as modifiers of genes whose protein products interact with the causal gene-encoded proteins in whole blood. Notably, we identified a causal association with HDL cholesterol through the extended networks in whole blood (Figure 3; Supplementary table 7a). However, since the regulatory effects of the SNPs are not restricted to whole blood, it is possible that the HDL – CAD linkage occurs in other tissues where it can mechanistically modify the CAD trajectory.

64 traits were significantly enriched within the CAD-associated network (Figure 3b; Supplementary table 7b). 14 of the 64 traits were associated with immune system traits (e.g. complement C4 levels and Type 1 diabetes; Figure 3b). A subset of regulatory loci associated with traits present in both the CAD-associated network level also regulate CAD-causal genes (Figure 3a – grey box). The majority of these shared traits were either CAD or CAD-proxy, or cholesterol traits (e.g. HDL and LDL cholesterol levels). Furthermore, 19 of the 64 traits enriched within the CAD-associated network were also present within the CAD-causal network (Figure 3a). Collectively, these observations are consistent with convergence between the causal and associated networks.

We hypothesised that the trait level convergence we observed between the traits in the CAD-associated and causal networks may extend to genetic convergence between the CAD-associated and CAD causal networks. Therefore, we tested to see if the genes that are within the CAD-associated network overlapped the 93 causal whole blood genes from the MR analysis in Europeans (Figure 1b). Of the 54 genes in the associated network (Figure 3a), forty-one genes were found to overlap with the set of 225 European causal genes for CAD within whole blood. Using the hypergeometric test, the observed overlap of 41 genes is significantly higher than an expected 0.623 genes, supporting our hypothesis.

### Protein interaction and gene regulatory networks associated with SARS-CoV-2 infection severity contain CAD causal genes

Protein interactions mediate fundamental biological processes enabling the functioning and regulation of all living organisms. We hypothesised that genes that modulate SARS-CoV-2 infection severity and causal CAD genes interacting closely within biological networks will have correlated effects^16,17^.

CAD was previously genetically associated with SARS-CoV-2 infection severity using tissue-specific protein-protein interaction networks^21^. Here we considered the hypothesis that epidemiological associations with CAD observed in the post-acute phase arise from a convergence between variants that affect SARS-CoV-2 infection severity and genes that are causal for CAD risk. Therefore, we tested the SARS-CoV- 2/CAD networks for the presence of the CAD causal genes (Figure 4a). We identified that genes causally associated with CAD significantly (hypergeometric *p* < 0.05) overlap with genes within the associated SARS-CoV-2 protein interaction network (Figure 4b and c). The causal CAD genes in the lung, blood and coronary artery SARS-CoV-2/CAD interaction networks across both ancestries overlap between the two SARS-CoV-2 phenotypes studied (hospitalised and severe; Figure 4b and c), however the proportions were not significant (coupon *p* > 0.05). The significance of the causal CAD nodes within the SARS-CoV-2 networks and the gene sets overlapping between phenotypes suggests these causal associations are independent of SARS-CoV-2 severity. These findings are consistent with epidemiological associations for cardiovascular disease in the post-acute phase of SARS-CoV-2, which occurs as frequently in mild cases of the infection as it does in severe cases^49–56^.

**Figure 4.**
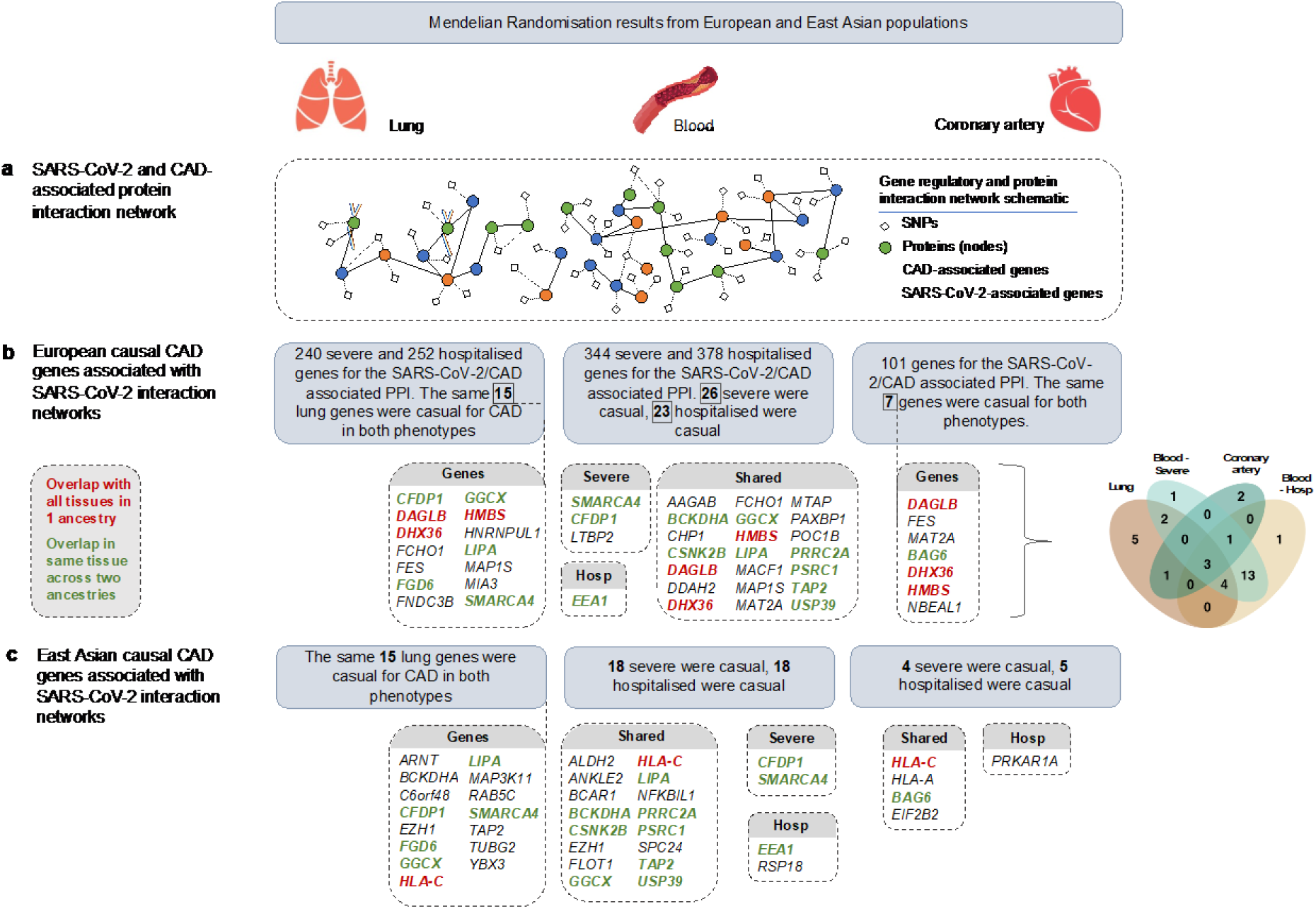
Genes within SARS-CoV-2 protein interaction networks overlap with genes causal for CAD. **(a)** We previously integrated protein-interaction networks with gene regulatory networks to identify networks of proteins and eQTLs that are associated with both SARS-CoV-2 and CAD^21^. These SARS-CoV-2/CAD specific PPIs were checked for causal nodes from the lung, whole blood, and coronary artery causal gene sets in European **(b)** and East Asian **(c)**. Causal genes present in all three tissues in European or East Asian ancestry are highlighted in red. Causal genes that overlap in the same tissue across European and East Asian ancestries are highlighted in green (inset).

## DISCUSSION

We identified a total of 98 novel protein-coding genes that are causal for CAD across coronary artery, whole blood and lung tissues, within European and East Asian ancestries. We identified a further 152 genes, which had previously been identified as being causal for CAD by Costanzo et al.^38^, supporting our approach. Protein interaction networks enabled us to identify pathways involving causal genes that are genetically associated with known co-occurring traits such as HDL cholesterol, that had previously not been causally linked with CAD. Thus, our findings extend on the knowledge of CAD risk from GWAS studies^5,6^, highlighting causal mechanisms that are tissue-agnostic and ancestrally unique without *a priori* assumptions.

Integrating protein interactions into our de novo comorbidity pipeline identified CAD associations with cholesterol. While genotypes increasing LDL cholesterol are known to elevate CAD risk^48^, a causal link with HDL-C cholesterol^57^ has been elusive^58^. We discovered modifiers of causal-CAD genes connected to HDL-C metabolism. Notably, *ZNF664, UBASH3B, MARCH2, R3HDM2 and MAP3K11* were identified as causal for CAD, suggesting a potential causal link between HDL-C and CAD in whole blood.

Causal genes identified, across both ancestries, were involved in gene regulation or the processes associated with gene regulation. For example, *PAXBP*, is involved in DNA binding and transcriptional regulation. *KIAA0232*, identified as causal in East Asians, encodes a nuclear phosphoserine protein that is involved in gene expression and cell signalling ^59^. Within the coronary artery in Europeans, we identified causal genes, *USP32* and *APPBP2*, that are involved in the ubiquitin-proteasome system, which is a pervasive post-translational protein modification system^60^. Dysfunction of the ubiquitin-proteasome system has been associated with cardiovascular pathologies (eg atherosclerosis)^61^. *USP32* is a deubiquitinase, and *APPBP2* encodes a protein with ubiquitin ligase-substrate adaptor activity. In the lung in Europeans the causal gene, *CAMK1D* is part of kinase signalling, which has been associated with cardiac hypertrophy^62^.

CAD is a multi-system disease^57^. Therefore, multi-tissue approaches are required to understand CAD development, disease progression from adolescence, and linkages to metabolic disturbances^12^ and the immune system^13^. We contend that the genes we identified as being causal, across multiple tissues, are more likely to be involved in disease development and progression, and not severity per se. For example, *CFDP1* was identified as a CAD causal gene in lung and whole blood in Europeans and East Asians. *CFPD1* has previously been associated with CAD by GWAS in European^63^ and East Asian^9–11^ cohorts. *CFPD1* has also been associated with blood pressure, aortic diameter, neuroblastoma susceptibility and carotid intima–media thickness^63–66^ in European cohorts.

Here, we tested all variants within the GTEx dataset without *a priori* assumptions using GRNs to assess those with putative regulatory functions for MR in CAD. We identified a novel variant (rs9934007) in the *BCAR1-CFDP1-TMEM170A* locus, as being associated with the regulation of the expression of *CFDP1*^37^. We identified this variant (rs9934007) as being protective for CAD in whole blood and lung in both Europeans and East Asians. In addition, we identified the novel variant rs11645691 in the *BCAR1- CFDP1-TMEM170A* locus as being causally associated with an increased risk of CAD in the coronary artery in East Asians. We contend that the novel causal association of rs11645691-*CFDP1* contributes to the genetic architecture of CAD in East Asians in the coronary artery.

We present evidence that alterations in gene isoform transcript levels may contribute information about the mechanisms through which genes are causal, by identifying isoforms whose transcript levels correlate with protection. For instance, *RPS18*, has a reduction in protein-coding isoforms that appear to associate with a shift from risk to protection between different tissues (whole blood and coronary artery). Arguably, treatment to replace the coding transcript represents a potential therapeutic approach to reduce CAD risk. By contrast, for *MAT2A*, we identified an increase in expression of isoforms that associated with risk within the coronary artery (compared to whole blood). We contend that the potential to identify causal isoforms and not simply genes, highlights the benefit of a multi-tissue MR approach for CAD causality.

This study is not without limitations and study bias, which arises from our reliance on existing Hi-C and GWAS datasets. As mentioned earlier, to be included in MR, an eQTL variant must (i) exhibit strong associations with the exposure, (ii) be associated with the outcome through the tested exposure only, and (iii) have no associations with any potential confounders that affect the outcome. The methods we employed minimised the possibility of violating these assumptions. However, it remains possible that this is not true during all developmental windows, as expression and genome organization change during development. In addition, our analyses exclude spatial eQTL target genes that lack curated interactions in resources like STRING or PROPER, potentially overlooking valuable insights. Another limitation pertains to the static nature of the gene regulatory and protein interaction networks we constructed, offering only a snapshot of potential interactions that may evolve over time. Lastly, our analyses amalgamate data from diverse sources (eg eQTL data from GTEx, Hi-C datasets, GWAS, and protein interaction data). This lack of overlap is a strength that enables the integration of the GRNs with the GWSS data. However, it can be considered to weaken the GRNs themselves due to the fact that the genome organization and eQTL data are not from the same biological samples. Despite these limitations, our approach enhances our ability to discern the direct contributions of causal genes and associated traits in CAD.

## Conclusion

98 genes are causally associated with CAD in a ancestry-specific manner that is consistent with the existence of a shared mechanism and specific gene x environment contributions to CAD. Specific gene isoforms, and not simply total expression, may contribute to the causal relationships. Finally, there is evidence for a causal connection between HDL cholestrol and CAD mediated through *ZNF664, UBASH3B, MARCH2, R3HDM2 and MAP3K11*. Future studies should confirm the functional impacts of the sceQTLs and the causal contributions of the genes that we identified.

## Data Availability

All data produced in the present work are avialble at:
European GWAS Data for CAD: https://gwas.mrcieu.ac.uk/datasets/ebi-a-GCST005195/
East Asian GWAS Data for CAD: https://gwas.mrcieu.ac.uk/datasets/bbj-a-159
Two-Sample MR package: https://mrcieu.github.io/TwoSampleMR/

https://gwas.mrcieu.ac.uk/datasets/ebi-a-GCST005195/

https://gwas.mrcieu.ac.uk/datasets/bbj-a-159

https://mrcieu.github.io/TwoSampleMR/

## Acknowledgements

The authors would like to thank the Liggins Systems biology group for useful discussions.

The Genotype-Tissue Expression (GTEx) Project was supported by the Common Fund of the Office of the Director of the National Institutes of Health and by NCI, NHGRI, NHLBI, NIDA, NIMH, and NINDS. The authors thank the Genomics and Systems Biology Group (Liggins Institute, University of Auckland) for valuable discussions. The authors acknowledge the COVID-19 Host Genetics Initiative consortium for providing infrastructure and access to the SARS- CoV-2 GWAS meta- analysis data.

## Availability of data and materials

European GWAS Data for CAD: https://gwas.mrcieu.ac.uk/datasets/ebi-a-GCST005195/

East Asian GWAS Data for CAD: https://gwas.mrcieu.ac.uk/datasets/bbj-a-159 Two-Sample MR package: https://mrcieu.github.io/TwoSampleMR/

## Competing interests

The authors declare no competing interests.

## Ethics

This project was performed under approval from the University of Auckland Human Participants Ethics Committee (project number UAHPEC-020905; Decoding SNPs in context).

## Funding

The Sir Colin Giltrap Liggins Institute Scholarship funded RKJ. The Dines Family Charitable Trust funds JOS.

## Author contributions

RKJ performed data analyses and interpretation, created figures, and wrote the manuscript. JOS directed the study, contributed to data interpretation, and co-wrote the manuscript.

## METHODS

### Generation of gene regulatory networks

We generated gene regulatory networks (GRNs), which included all spatially constrained eQTLs for all known SNPs (MAF ≥ 0.05; dbSNP154^67^) for lung, whole blood (dbGaP accession: phs000424.v8.p2; approved project number: #22937) and coronary artery (GTEx v 8.0)^37^. SNPs were screened through CoDes3D, one chromosome at a time. Multiple testing was corrected using the Benjamini-Hochberg procedure (FDR ≤ 0.05) and interactions were kept if the logarithm of allelic fold change (log_aFC) ≥ 0.05^68^.

### Identification of SNP-gene pairs following infection with SARS-CoV-2

Genome-wide association study (GWAS) data for SARS-CoV-2 clinical phenotypes was obtained from the Covid-19 Host Genetics initiative^69,70^ COVID-19 HGI). Single nucleotide polymorphisms (SNPs) for the hospitalised versus population and severe (hospitalised AND death or respiratory support) versus population (*p*-value threshold of 1 x 10^-5^) cohorts were obtained from COVID-19 HGI release 7 (https://www.covid19hg.org/results/r7/). SNPs in linkage disequilibrium within 5,000 base pairs at an R^2^ threshold of > 0.8 from European populations were obtained from 1000 Genomes phase 3 data^71^. Full summary statistics and details from COVID-19 HGI are available at https://app.covid19hg.org/44.

### Assigning putative transcriptional functions to SARS-CoV-2 SNPs

Severe and hospitalised SARS-CoV-2 associated SNPs were analysed separately using CoDes3D^68^ to identify phenotype-specific spatially constrained expression quantitative trait loci (sceQTLs) and their target genes (Supplementary tables 4.5a-f). Phenotype-specific (i.e. hospitalised or severe) spatial connections for each SNP-gene pair were identified from Hi-C chromatin contact data derived from human lung primary tissue^72^, blood (peripheral blood B cells, peripheral blood CD4^+^ T cells, peripheral blood CD8+ T cells^39^, peripheral blood T cells^73^), and the coronary artery (smooth muscle cells^74^). To identify which SNPs are eQTLs, the SNP-gene pairs were used to query lung, whole blood, brain cortex and the coronary artery within the GTEx database^37^. Multiple testing was corrected using the Benjamini-Hochberg procedure (FDR < 0.05) and interactions were kept if the logarithm of allelic fold change (log_aFC) ≥ 0.05^68^. eQTL and gene chromosome positions were annotated according to human reference genome GRCh38/hg19.

### Gene set enrichment analysis (GSEA) in tissues closely related to whole blood

Causal genes associated with CAD in blood tissue in the European population were screened for gene set enrichment^40,41^ in artery plaque samples from CAD patients with stable and advanced disease^42^. GSEA evaluates the functional significance of gene expression data (RNA-seq) by incorporating our predefined gene set (93 causal blood genes) and testing for whether they exhibit statistically significant enrichment or depletion in the artery plaque samples^42^. Briefly, GSEA ranks all genes within the expression dataset based on differential expression between the stable and advanced CAD patient samples. The ranking was conducted using signal-to-noise ratios. Enrichment score (ES) is then calculated by analysing the distribution of ranked genes within the gene set. The ES reflects the degree to which the genes in the set are overrepresented at the top or bottom of the ranked list. A permutation-based test (n = 1000) was performed to determine if the observed enrichment score was statistically significant. It repeatedly shuffles the gene labels in the dataset, recalculates the enrichment scores for each permutation, and generates a null distribution of scores. Multiple testing was corrected using the Benjamini-Hochberg procedure (FDR < 0.05) to control for false positives.

### Identification of causal nodes within SARS-CoV-2/CAD protein interaction networks

All genes contained within previously identified SARS-CoV-2/CAD protein and gene regulatory interaction networks were screened for the tissue-specific causal CAD genes we identified here. The screening was done backwards based on the causal CAD genes. For example, Where Gene_c_ was associated with CAD, Gene_b_ and Gene_a_ were obtained from the overall SARS-CoV-2 interaction network (interactions Gene_a_ – Gene_b_ – Gene_c_; FDR *p* < 1 x 10^-5^). Reconstructed protein interaction networks where a causal node was identified were visualised in Cytoscape and screened for validated interactions and pathway analysis using the STRING^75,76^ protein interaction tools (with all interaction sources and high confidence > 0.7). Testing for the significance of the overlap between CAD-associated and CAD-causal network genes was conducted using the hypergeometric test. Given:

- Total number of genes N = 19,500 (protein-coding genes in the human genome{Amaral, 2023 #883})
- Number of genes in the European causal genes for CAD within whole blood K = 225
- Number of genes in the CAD-associated network n = 54 The expected number of overlapping genes can be calculated as:

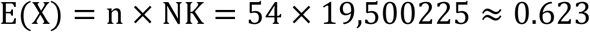

### Mendelian randomisation

We previously produced^77^ a lung gene regulatory network (GRN) consisting of 908,356 spatial eQTLs (731,067 SNPs [MAF ≥ 0.05] and 15,532 genes) that were identified within lung tissue (GTEx^37^). The whole blood GRN previously generated^33^ consisted of 1,634,655 eQTLs (720,675 SNPs [MAF ≥ 0.05] and 14,556 genes). The coronary artery GRN consisted of 490,215 eQTLs (144,399 SNPs [MAF ≥ 0.05] and 6,542 genes). To identify the independent tissue-specific eQTLs as instrumental variables, we filtered each GRN for eQTLs with a *p* < 1 x 10^-5^ and performed LD clumping. Using the “clumping” function within the ‘TwoSampleMR’ package, this was achieved using PLINK (v1.90b6.21) with an r^2^ < 0.001, 10Mb away with a European population reference. The remaining SNPs were used as genetic instruments for testing causal GRN genes associated with coronary artery disease.

Two-sample mendelian randomisation was conducted using the R package TwoSampleMR. We used fixed-effects, inverse-variance-weighted MR for proposed instruments that contain more than one variant and Wald ratio for proposed instruments with one variant. For proposed instruments with multiple variants, we also tested the heterogeneity across variant-level MR estimates, using the Cochrane Q method (mr_heterogeneity option in TwoSampleMR package). We defined significant MR results using a P value threshold of p < 1 x 10^-5^. Causal genes were checked for overlaps within the Cardiovascular Disease Knowledge Portal^38^, which aggregates and analyses genetic association results, epigenomic annotations, and results of computational prediction methods to provide data, visualizations, and tools in an open-access portal.

There is no overalp between the individuals the GWAS^5,6,36^ or GTEx study (GTEx v 8.0)^37^, as they were independent, and have not been merged.

## Notes

### Competing Interest Statement

The authors have declared no competing interest.

### Author Declarations

This study used only openly available human data that were orginally located at:European GWAS Data for CAD: https://gwas.mrcieu.ac.uk/datasets/ebi-a-GCST005195/ East Asian GWAS Data for CAD: https://gwas.mrcieu.ac.uk/datasets/bbj-a-159

